# Health care workers’ self-perceived infection risk and COVID-19 vaccine uptake: a mixed methods study

**DOI:** 10.1101/2022.10.07.22280829

**Authors:** Kasusu Nyamuryekung’e, Maryam Amour, Innocent Mboya, Harrieth Ndumwa, James Kengia, Belinda J Njiro, Lwidiko Mhamilawa, Elizabeth Shayo, Frida Ngalesoni, Ntuli Kapologwe, Albino Kalolo, Emmy Metta, Sia Msuya

## Abstract

**Background:** Vaccination is the most cost-effective way of preventing COVID-19 disease although there was a considerable delay in its institution in Tanzania. This study aimed to assess healthcare workers’ (HCWs) self-perceived infection risk and uptake of COVID-19 vaccines.

**Methods:** A concurrent embedded, mixed methods research design was utilized to collect data among HCWs in seven Tanzanian regions. Quantitative data was collected using a validated, pre-piloted, interviewer administered questionnaire whereas in-depth interviews and focus group discussions gathered qualitative data. Descriptive analyses were performed, and chi-square test used to test for associations across categories. Thematic analysis was used to analyze the qualitative data.

**Results:** A total of 1,386 HCWs responded to the quantitative tool, 26 participated in the in-depth interviews and 74 in the focus-group discussions. About half of the HCW (53.6%) reported to have been vaccinated and three quarters (75.5%) self-perceived to be at a high risk of acquiring COVID-19 infection. Participants perceived that the nature of their work and the working environment in the health facilities increases their risk to infection. Limited availability and use of personal protective equipment was reported to elevate the perceived risks to the infection. Respondents belonging in the oldest age group and from low and mid-level health care facilities had higher proportions with a high-risk perception of acquiring COVID-19 infection compared to their counterparts.

**Conclusions:** Only about half of the HCWs reported to be vaccinated albeit the majority recounted higher perception of risk to contracting COVID-19 due to their working environment, including limited availability and use of personal protective equipment. Efforts to address heightened perceived-risks should include improving the working environment, availability of PPEs and continue updating HCWs on the benefits of COVID-19 vaccine to limit their risks to the infection and consequent transmission to their patients and public.

## Introduction

The novel coronavirus disease 2019 (COVID-19) caused by severe acute respiratory syndrome corona virus 2 (SARs-CoV-2) remains a significant disease of public health concern. Since its emergence, it has been shown to spread rapidly causing dramatic global health crisis [1]. It was declared as a pandemic by the World Health Organization (WHO) on the 11th of March 2020, with a total of 56,837,067 confirmed cases, 340,447 new cases and 928,593 confirmed deaths globally as of January 17, 2022 [2]. Tanzania had reported relatively few number of COVID 19 cases, with a total of 31,395 confirmed cases and 745 deaths reported between January 2020 and January 18, 2022 [3].

Vaccination is among the most cost-effective ways of preventing diseases. For the vaccination effect to be appreciated, several strategies to be considered include vaccine availability, accessibility, acceptability, and willingness of the population to vaccinate. Studies show that about 14.3 % and 22.1% of the global population intend to refuse vaccination or showed uncertainty respectively, with higher rates reported in lower income countries [3]. Moreover, perceived vaccine efficacy and safety concerns contribute to the observed trends in most countries [4].

There was a considerable delay in COVID 19 vaccine roll out in Tanzania, after launching the first nation-wise COVID 19 vaccination, the first roll out was made available among priority groups; health care workers (HCWs) with high risk of getting and transmitting the infection, people with advanced age and underlying medical conditions with a high risk of developing severe disease [5]. A total of 2,431,769 vaccine doses administered by the end of year 2021 under COVAX facility [3,6]. While vaccine availability may not have been a challenging option initially, the issue of vaccines acceptance and hesitancy influenced by social, political, and religious factors may contribute to low vaccine uptake.

HCWs stand as among the most important groups as trusted influencers in regards to health issues, including vaccination decisions [7]. This important group should be guided and supported to provide credible and scientifically proven information on vaccines as their influence in the community remains pivotal. It is therefore important to understand and acknowledge HCWs perspectives with regard to COVID 19 vaccines [4]. However, studies have reported a number of challenges facing this population including high risk of infection, insufficient personal protective equipment (PPE), heavy workloads and discrimination [8].

Risk perception, defined as an individual perceived susceptibility to threat, plays a key role in health behavioral change theories, including health decision making process [9]. HCWs are among the most vulnerable groups for SARs-CoV-2 infection, they work in frontline positions with suspected and confirmed COVID 19 cases, 5-7.3% of HCWs were found to be COVID 19 positive in some developed countries [10,11]. While being a high-risk group, some studies have reported that perceived risk of COVID 19 infection and detrimental health effects among HCW are associated positive protective behaviors [12]. In Ethiopia, 88% of HCWs were reported to perceive their risk of being infected with COVID 19 infection as high, and showed widespread practice on preventive measures [13].

Myths and misconceptions around the COVID 19 vaccine subject have been circulating and its impact can be observed especially in developing countries [1]. This has been shown to contribute to the observed vaccine hesitancy, which is defined by WHO as the reluctance in accepting vaccines or an outright refusal of vaccines despite their availability [7]. WHO has further mentioned vaccine hesitancy as one of the top global threats to public health in 2019 [7,14]. As reported by a study done in Senegal with 5.5% COVID 19 vaccine coverage, vaccine hesitancy and refusal have contributed to low vaccine uptake despite its multifaceted nature [15]. In Ethiopia, more than 50% of HCWs were found to be vaccine hesitant [4].

While being at an increased risk of COVID 19 infection and disease transmission in Tanzania, HCWs play an influential role in community understanding and overall vaccine uptake; there is paucity of data on the status of vaccine uptake among HCWs and the influencing factors in Tanzania. Understanding HCWs risk perception and their influence on vaccination is crucial in informing policy makers and highlighting educational needs to address the situation especially in developing countries like Tanzania. This mixed method study illustrates on HCWs perceptions in relation to the COVID 19 vaccine uptake situation in Tanzania.

## Materials and Methods

### Study setting

Tanzania is a large East African country with an area of 947,000 square kilometers and an estimated population of 61.5 million (2021 World Bank projections) of which about two-thirds live in rural areas. Tanzania comprises of the much larger mainland and semi-autonomous isles (Zanzibar); the current study was conducted in mainland Tanzania. Mainland Tanzania is administratively divided into 26 regions, each region comprising of a variable number of districts (4 – 6), which in turn contain wards. The Tanzanian healthcare delivery facilities follow this pyramidal administrative arrangement, with regional referral hospitals situated at the apex, functioning as the highest-level hospital within a region, which receive referrals from district hospitals, that in turn receive patients from lower levels (health centers and dispensaries). Groups of regions are further clustered into six geographical zones (Northern, Coastal, Central, Southern highlands, Western and Lake). One region was randomly selected from each of these six zones to be included in the study. The sampled regions were Kilimanjaro, Lindi, Njombe, Mbeya, Tabora and Simiyu representing Northern, Coastal, Central, Southern highlands, Western and Lake zones, respectively. Dar es salaam as a cosmopolitan and the largest city in Tanzania was also purposefully selected to be included in the study.

### Study design and participants

A concurrent embedded mixed methods research design was utilized to collect data among HCWs in seven regions of mainland Tanzania from November 2021 to January 2022. The qualitative part of the study was embedded in the quantitative cross-sectional study. The qualitative part was mainly intended to explain the healthcare workers’ risk perceptions towards COVID-19 disease, as a supplement of the quantitative study assessing COVID-19 vaccine hesitancy.

A sample size for the quantitative part of the study (N = 1400) was determined by using a single proportion formula taking a standard normal value of 1.96 under the 95% confidence limit, 50% proportion of vaccine hesitancy (for maximization of sample size), 3.5% margin of error, 1.5 design effect to address the clustering effect while adjusting for a non-response rate of 20%.

Multi-stage sampling technique was employed to recruit HCWs from the seven (7) selected regions for the quantitative part. One Regional Referral hospital, two (2) district hospitals and two (2) health centers from each of the identified regions were included in this study. Therefore, a total of seven (7) Regional Referral Hospitals, fourteen (14) district hospitals, and fourteen (14) health centers were included. Systematic sampling technique was used to select healthcare facilities for inclusion. Sampling of HCWs within the selected health facilities was based on their number in the selected health facilities in a region proportional to their size. Upon determination of the respective health facilities’ sample sizes, HCWs were consecutively invited to participate and enrolled into the study.

The qualitative component of the study was conducted in four of the seven regions where the quantitative study took place. In-depth interviews were conducted in each region with the key officials including the Regional Medical Officers, Regional Vaccination Officers, District Medical officers, District vaccination officers and hospital in charges leading to a total of 26 interviews. Additionally, we conducted two focus group discussions in each region with participants ranging from 6 – 12 people leading to a total 74 participants in 8 FGDs. The FGDs engaged health care workers in the selected districts within the study regions.

### Data collection tools and procedures

Quantitative data was collected using a validated, pre-piloted questionnaire through the Open Data Kit (ODK). The questionnaires were developed based on various studies and WHO proposed questions to assess vaccine hesitancy and acceptability [16–21]. The questionnaire was prepared in English and translated in Swahili and had four components: socio-demographic, awareness and knowledge on COVID-19 vaccines, risk perception towards COVID-19 and COVID-19 vaccine acceptance. Back translation to English was done to preserve the meaning of the questions. The questionnaire was administered face-face by trained research assistants (RAs). On the day of quantitative data collection, the RAs visited the HCWs, introduced themselves and explained the study purpose. Then, consent information was administered in a quiet, private place around the health facility. Special emphasis was placed on issues of anonymity and confidentiality, and in assuring the respondents that no personal identifiable information will be collected to encourage truthful responses. Only the consenting individuals were interviewed.

Qualitative data was collected through IDIs and FGDs with purposively selected health officials and HCWs to explore their opinions and risk perceptions towards COVID-19. All interviews were conducted in Swahili and audio recorded with the permission of the study participants. Further, researchers applied the principle of bracketing to ensure that pre-understanding information do not influence the data [22]. Furthermore, for enhancement of reliability, field notes as a reflective diary were maintained.

### Data management and analysis

The collected quantitative data was transferred from the Open Data Kit (ODK) to an excel spreadsheet. Upon completion of data collection, each questionnaire was assessed for its completeness. Data entry, cleaning and coding was done using Microsoft Excel program and exported to Stata software V.16.1 (College Station, Texas). Descriptive analyses were performed for proportions, percentages, means and their corresponding standard deviations.

The primary outcome variable of the study was COVID-19 risk perception which was assessed by asking a question “How do you perceive the level of risk that you have for acquiring COVID-19 infection” with responses along a six-point Likert scale ranging from “Not at all” to “Very high risk”. Thereafter, the responses were dichotomized into a “Low risk” and “High risk”. Vaccination status of the respondents was assessed by asking “Have you been vaccinated against COVID-19” with “Yes/No” responses.

Age and work experience were recoded into categorical variables. For categorical variables chi-square test was used to assess associations between sociodemographic characteristics and COVID-19 vaccination status to Risk perceptions. Statistical significance was defined as a p value of <0.05.

For qualitative data, the audio recorded in-depth interviews and focused group discussions were transcribed verbatim into word file documents where non-verbal cues were also considered. The transcription process started within 24 hours after the conduct of the interview to allow follow-up on issues for more clarity and determination of data saturation in subsequent interviews and discussions. The transcribed transcripts were checked against the audio records by two of the research team members to ensure accuracy and quality of the data generated. Thematic analysis was used to analyze the information following the five stages as described by Braun and Clarke, 2014 (23) to establish meaningful patterns in the data: familiarization with the data, generating initial codes, searching for themes among codes, reviewing themes and presenting the results. The coding also involved identification of the typical quotes that are used to illustrate the various themes presented in the study.

### Ethical considerations

Ethical approval was obtained from the Research and Publication Committee of the Muhimbili University of Health and Allied Sciences (MUHAS-REC-08-2021-839). Permission to collect data in Regions and Councils was sought from the President’s Office Regional Administration and Local Government, Ministry of Health Community, Development, Gender, Elderly and Children (MoHCDGEC), Regional Secretariat (RS) and Local Government Authorities (LGAs). Prior to collection of data, all participants were provided with information on the purpose of the study, voluntary nature of participation, right to withdraw from study at any time without consequence and guaranteed anonymity. Signed, informed consent was obtained from all participants before enrolment into the study.

## Results

A total of 1368 health care workers were approached and involved in the quantitative part of this study. Most of the respondents were female (60.1%) and had the mean age of 35.7 years (SD 10.1). There was almost an equal representation of participants by regions, except for Dar es Salaam which contributed the largest proportion (26.1%). Most of the respondents were from the district-level facilities (42.1%) and about three quarters (77.5%) worked in Government facilities (Table 1).

**Table 1:**
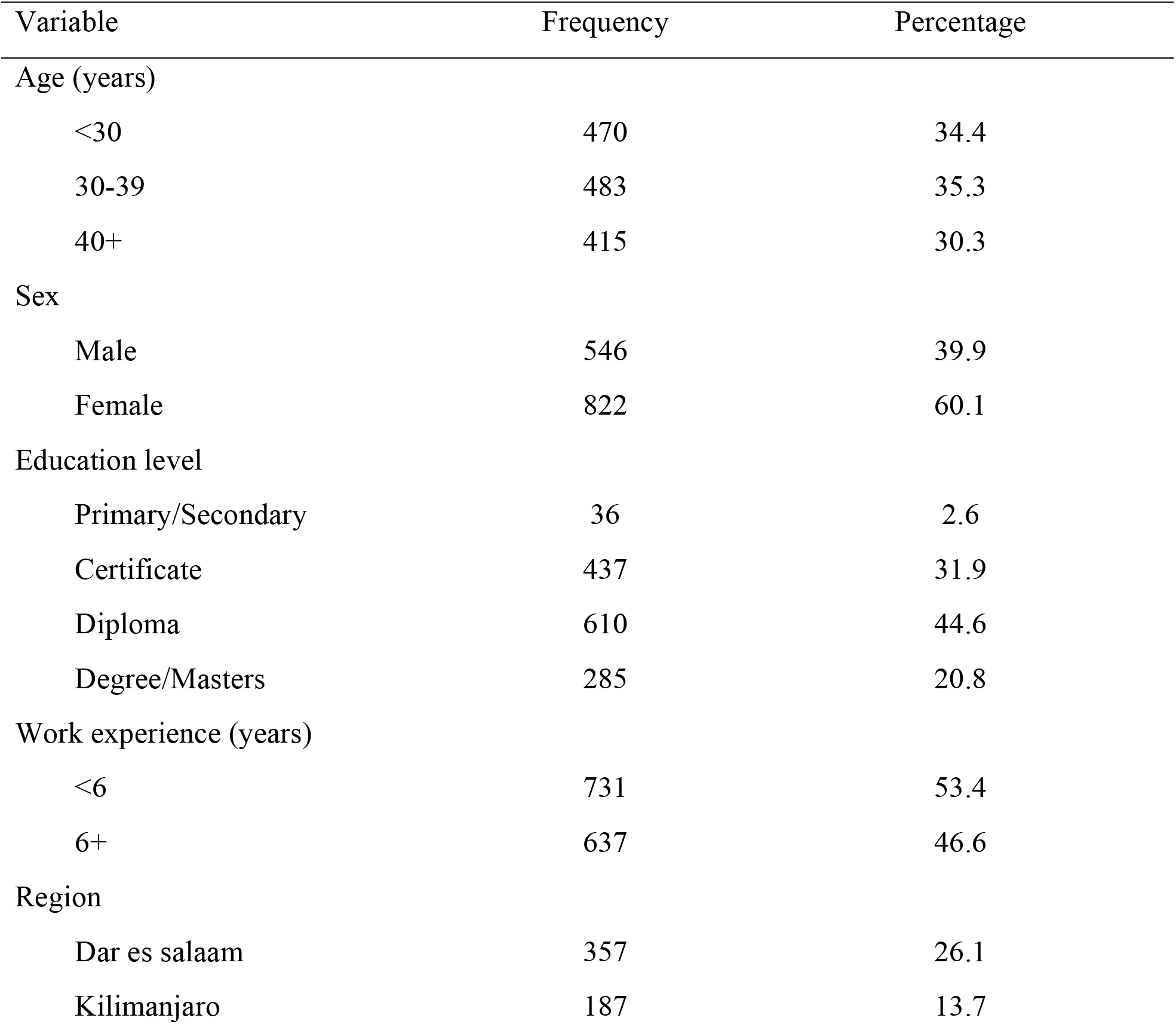
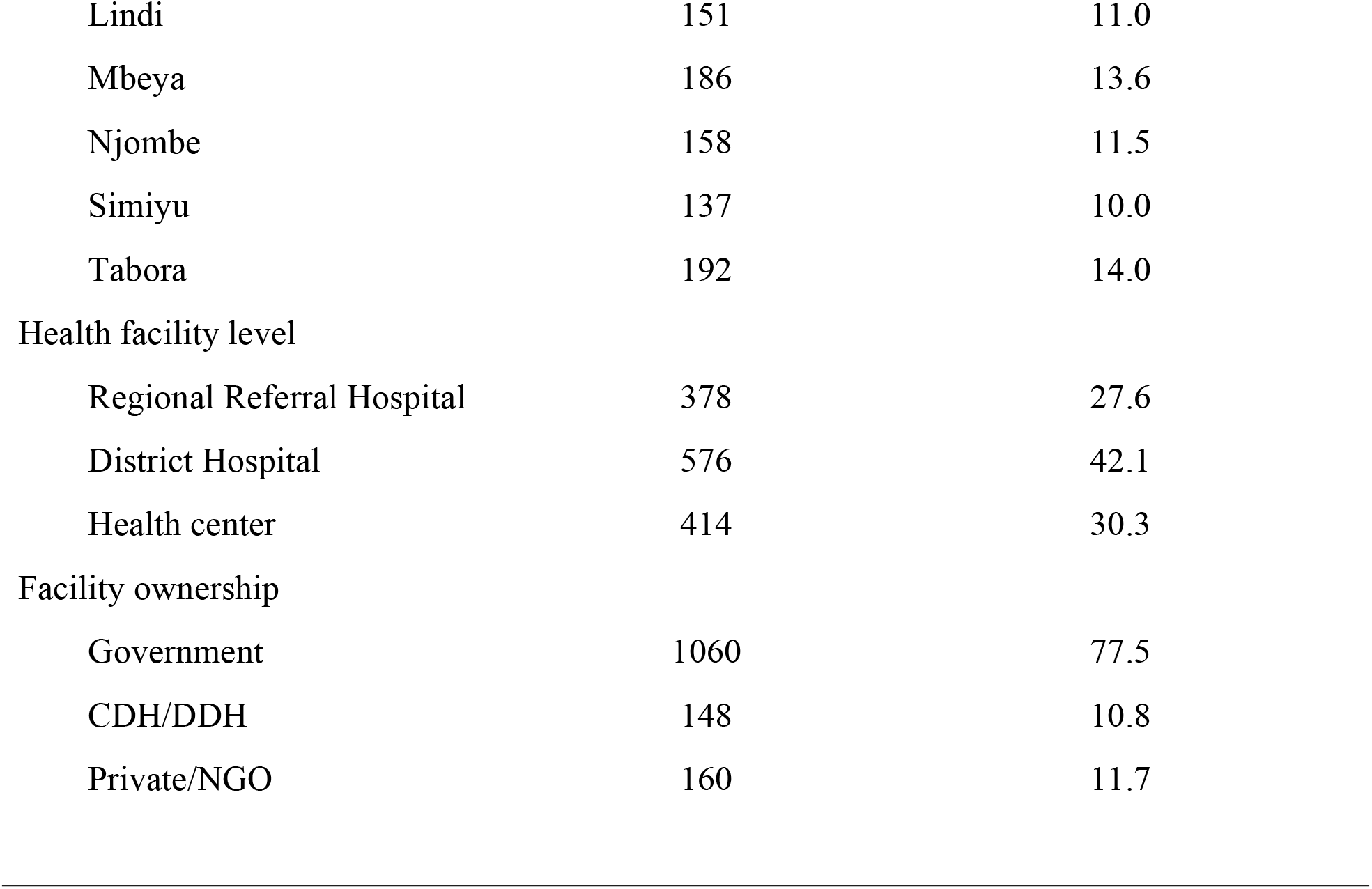
Background characteristics of HCWs (N=1368)

Only about one half of the HCW (53.6%) reported to have been vaccinated whereas three quarters (75.5%) self-perceived to have a high risk of acquiring a COVID-19 infection. Accordingly, those that perceived to have a high risk for COVID-19 infection had a larger proportion reporting to have been vaccinated for COVID-19 compared to their counterpart (p <0.01) (Table 2).

**Table 2:**
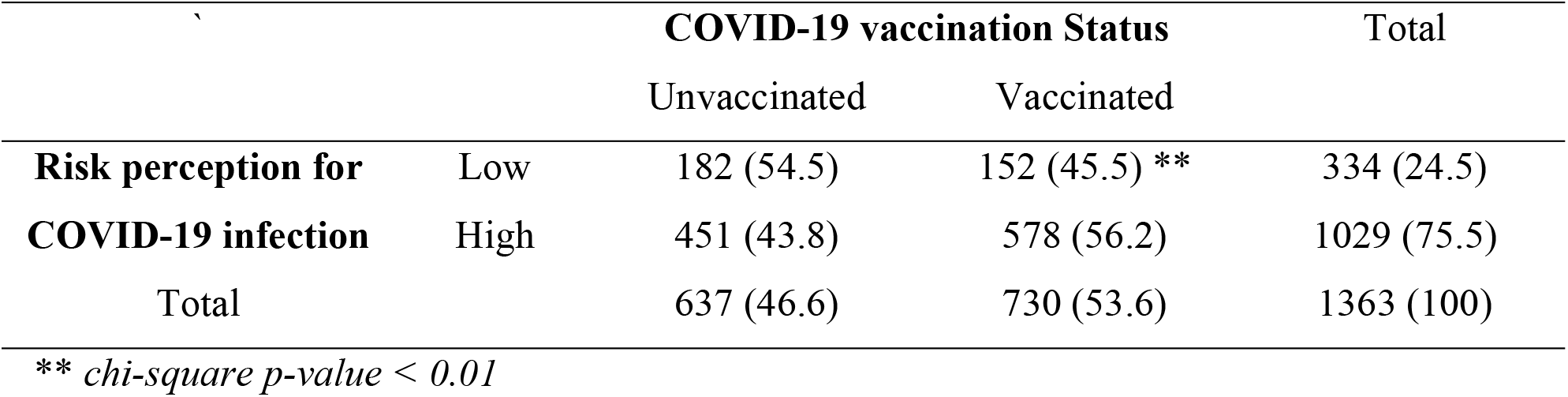
Vaccination status and risk perception for COVID-19 infection

**Table 3:** Socio-demographic characteristics by high COVID-19 risk perception and reporting being vaccinated for COVID-19

| Variable | High Risk Perception | p-value | Vaccinated | p-value |
| --- | --- | --- | --- | --- |
| **Age (years)** |  |  |  |  |
| <30 | 335 (71.4) |  | 187 (39.8) |  |
| 30-39 | 350 (72.8) | .000 | 271 (56.1) | .000 |
| 40+ | 344 (83.3) |  | 273 (65.8) |  |
| **Sex** |  |  |  |  |
| Male | 399 (73.5) | .159 | 292 (53.5) | 1.000 |
| Female | 630 (76.8) |  | 439 (53.4) |  |
| **Education level** |  |  |  |  |
| Primary/Secondary | 30 (83.3) |  | 20 (55.6) |  |
| Certificate | 344 (78.9) | .083 | 213 (48.7) | .121 |
| Diploma | 451 (74.4) |  | 337 (55.2) |  |
| Degree/Masters | 204 (71.6) |  | 161 (56.5) |  |
| **Work experience (years)** |  |  |  |  |
| <6 | 529 (72.8) | .012 | 319 (43.6) | .000 |
| 6+ | 500 (78.6) |  | 412 (64.7) |  |
| **Region** |  |  |  |  |
| Dar es salaam | 211 (59.3) |  | 177 (49.6) |  |
| Kilimanjaro | 146 (78.1) |  | 114 (61.0) |  |
| Lindi | 106 (70.2) | .000 | 93 (61.6) | .001 |
| Mbeya | 157 (85.3) |  | 87 (46.8) |  |
| Njombe | 142 (90.4) |  | 78 (49.4) |  |
| Simiyu | 114 (83.8) |  | 87 (63.5) |  |
| Tabora | 153 (79.7) |  | 95 (49.5) |  |
| **Health facility level** |  |  |  |  |
| Regional Referral Hospital | 262 (69.7) |  | 174 (46.0) |  |
| District Hospital | 454 (79.1) | .004 | 304 (52.8) | .000 |
| Health center | 313 (75.8) |  | 253 (61.1) |  |
| **Facility ownership** |  |  |  |  |
| Government | 777 (73.5) |  | 612 (57.7) |  |
| CDH/DDH | 121 (81.8) | .006 | 70 (47.3) | .000 |
| Private/NGO | 1313 (82.9) |  | 49 (30.6) |  |

The higher perceived risk to COVID 19 infection perceptions were also reported during the qualitative in-depth interviews and focus group discussions. When detailing the risks to COVID-19 infections participants voiced that the nature of the work done by HCWs and the working environment at the health facilities increases their risks to the infection. Limited availability and use of personal protective equipment’s (PPEs) including standard face masks and sanitizers at the health facilities were mentioned to elevate the perceived risks to the infection. Some HCWs reported that many times they must attend patients without using any PPEs because they are frequently unavailable. Others said that the risk to contacting COVID 19 infection is so high because even when the PPEs are available others HCWs do not comply with their use. One of the district officials when explaining about risks to COVID-19 infection he related it with the working environment as follows:

> *“the risk to COVID-19 infection among health care workers is high because of the working environment, people have relaxed, they are no longer taking measures against COVID 19, some do not bother to even wear mask, wash hands, keep social distancing and even when masks are there they just don’t put on all the time as required, everything about COVID-19 seem to paralyze, no one is either complying or discussing about it, which cause the working environment unsafe (IDI1)*.

Working in the health facility setting was reported as increasing the risk to contacting the COVID-19 infection. Participants voiced concerns that it is the HCWs who take care of the COVID-19 patients that also increase their likelihood of being infected. They said most of the hospitals for example do not have enough offices/ exchange rooms rather rooms are shared among HCWs including those that attend patients at the intensive care unit (ICU) or patients having trouble breathing. The shared rooms are small and limited in space a situation that increases their risk to COVID-19 infections. When detailing on this matter a participant during the in-depth interviews reported unless compliance to the recommend preventive measures is high, HCWs will continue to be at higher risk to contacting COVID-19:

> *“You cannot say health care workers are not at risk of COVID-19 as far as they are working in the hospital, they are taking care of the COVID-19 patients, they share small rooms, no dedicated rooms for those attending patients at the intensive care unit or with difficult breathing, sometimes do not have all the required PPEs so the risk is there and if one gets infected it is likely the rest will experience the same unless compliance to recommendation protective measures is high we will continue to be at higher risk of COVID-19” (IDI3)*

Participants believed the risk to COVID-19 infection is not only higher for HCWs but also increased the potential for them to transmit the infection to their patients. They said sometimes HCWs attend patients before even knowing either they have the infection or the patient they attend is having the infection due to the resemblances of COVID-19 symptoms with other diseases. This was elaborated during in-depth interviews by the health facility in-charge as follows:

“*Transmission of the infection is not avoidable … as long as you attend patients, a chance of acquiring or transmitting it to others still viable. First of all, you may not even be aware that you are COVID-19 positive when attending a patient because COVID symptoms resemble that of other diseases like malaria such as feeling fever, joint pains, cough and so on…and the patient may have same symptoms and you think it is malaria and not COVID-19*” (IDI4).

On the other hand, ideas that the risk to COVID-19 infection has been reduced with the use of COVID-19 vaccinations were also expressed. When explaining on this, participants compared the risk to the infection from the first wave when people did not know what to do about it with the time when vaccines were introduced:

“*The risk to COVID-19 infection was very high like 100% during the first wave because COVID 19 was a new thing and we had no enough knowledge on the precautions to take or what to do, but the risk decreased during the second wave because we had enough knowledge on how this disease is transmitted and how to take precautions and the risk decreased more and more to the point that we are not that much worried because there is the introduction of vaccine which has helped us to build protection” (FGD 3)*

Those respondents belonging in the oldest age group had higher proportions with a high-risk perception for acquiring COVID-19 infection compared to the younger age groups. Similarly, the proportion of respondents reporting to have been vaccinated for COVID-19 was highest within the oldest age group. Risk perception and vaccination status was also shown to vary significantly by region of the respondents. Whereas Njombe, Simiyu and Mbeya had more than 85% of the respondents perceiving their risk as high, those in Dar es Salaam only had about 60% reporting the same. On the other hand, respondents from Simiyu, Lindi and Kilimanjaro had more that 60% reporting to have been vaccinated compared to other regions which consistently had less that 50% reporting the same.

Respondents from low and mid-level of health care facilities (health centers and district hospitals, respectively) reported much higher risk perceptions compared to those in high level facilities (Regional referral hospitals). Equally, respondents in low level facilities had higher proportions reporting to have been vaccinated compared to those in high level facilities.

Respondents working in government facilities had a lower proportion (73.5%) reporting to be at high-risk compared to those working in faith-based organization, NGOs, and private health facilities. Contrariwise, those respondents from the government facilities had higher proportions reporting to have been vaccinated compared to their counterparts.

## Discussion

This study aimed at exploring HCWs perceptions in relation to the COVID 19 vaccine uptake to inform policy makers and highlighting targeted educational needs to address the similar situation especially in developing countries like Tanzania. About a quarter of the HCW perceived to have a low risk of acquiring a COVID-19 infection. Furthermore, those with perceived low risk had higher proportions reporting to be unvaccinated for COVID-19.

In the current study, majority of the HCWs perceived risk of contracting COVID-19 to be high, consistent to a recent multi-country study by Dryhust et al. that found equally high levels of COVID-19 risk perception levels in the countries [24]. Due to the nature of their daily work activities and physical proximity to potential COVID-19 cases, it was expected that the vast majority of the HCW in health facilities would consider themselves to be at a heightened risk for contracting the infection. However, consisted availability of appropriate and required PPEs would have contributed towards allaying some of the perceived risks. To control the spread of infection, it is crucial that all HCWs become sensitized to the increased risk that they are subjected to with respect to COVID-19 infection. This may ensure that necessary precautions and protective measures are adopted by the HCW and respective health facilities to prevent acquisition of infections, but more importantly, that they not become the source of infection to the patients and clients that they encounter regularly. High perceived risk of COVID-19 has largely improved the infection prevention and control behaviors of HCWs as indicated by studies in Egypt and Ethiopia [25] however, the picture was different in Tanzania where even though participants reported high perceived risk consistent use of protective gears was not reinforced, even when they were available.

It has been widely reported that high perceived risk of contracting COVID 19 is a significant predictor of vaccine acceptance. However, current findings reveal that only about a half of the HCW had been vaccinated-despite sustained efforts to ensure availability and encourage vaccinations. Thus, the link between the perceived risk of COVID-19 and COVID-19 vaccine uptake was tenuous in this setting, contrary to some similar studies [26]. One probable explanation for this could be that a significant proportion of people in Africa consider the vaccines as unnecessary, and that alternatives to COVID-19 vaccination exist [27]. Many studies have indicated that when people perceive COVID-19 as a threatening disease, the demand for a vaccine against the disease would be correspondingly increased. However, this study has shown that it is not necessarily the case and that other factors, especially vaccine safety concerns, might outweigh the perceived disease risks when an individual decides whether or not to accept the vaccine [28]. Informing the public about the safety of a COVID-19 vaccine should be the focus for health authorities aiming to achieve a high vaccine uptake especially in Tanzania where other factors including contradicting government stance may have had an influential role in overall vaccine acceptancy.

As expected, this study showed high perceived risk for COVID 19 among older HCW which correlated with high vaccine uptake. Literature indicate that vaccine hesitancy was more common among young people than older adults partly due to their lower risk of comorbidities [29–31]. Further, the observed excess mortality in the elderly population due to COVID-19 may have functioned to make this group feel particularly vulnerable, thus both enhancing their risk perception and increasing willingness to adopt protective measures. Being male has been reported to be uniformly associated with lower risk perceptions in many countries, which is consistent with other risk perception studies [32], a finding which was not corroborated in the present study. This may be due to the similarity of our participants with respect to the perceived risk of getting COVID-19 infection. That is, all HCWs have same risk for the infection, irrespective of their identified sex.

The triangulation method used in this study under mixed method design provides a deeper understanding and contextual insights of the research in question. The key informant interviews and the focused group discussions complement the quantitative findings. The possible limitation of conducting a mixed method study is the possibility of introducing Interview bias; to minimize this, in addition to training, authors provided a common interviewer guide to every interviewer.

## Conclusions

While majority of the HCWs perceived to have high risk of contracting COVID-19, only about a half of respondents reported to be vaccinated. Older age, female gender, working in a district hospital and a private owned hospital and high perceived risk for COVID-19 were associated with increased vaccine uptake. With the status of the working environment and constant exposure to patients, some HCWs perceived their risk of contracting COVID-19 to be unavoidable. However, other HCWs declared the potential role of COVID-19 vaccines in reducing their risk of infection. Targeted information the public on the beneficial role of COVID-19 vaccine in reducing transmission risk should be the focus for health authorities to achieve a high vaccine uptake in Tanzania.

## Policy implications

A consistent and evidence-based position adopted by the health authorities is an important prerequisite towards addressing any novel public health emergency. Tackling future public health emergencies requires deliberate actions to be taken in safeguarding the interests and wellbeing of the HCWs.

## Data Availability

All data and related metadata underlying the findings reported in a submitted manuscript should be deposited in an appropriate public repository, Dryad with the following DOI details: https://doi.org/10.5061/dryad.vdncjsxz9

## Acknowledgments

The authors acknowledge their corresponding institutions for providing support to conduct the study. We recognize the cooperation from all the health care workers that took part in this study. We acknowledge the support from the regional and district medical and vaccine officers and the health facilities-in charges of the involved health facilities. We thank our research assistants for their dedication to conduct this study timely, namely Melina Mgongo, Doris Mbata, Oko Okong’o, Zenaice Aloyce, Martha Joseph, Mtumwa Bakari, Nyanjura Manyama, Zenais Kiwale, Anastazia Ngowi, Barikiel Panga, Novatus Tesha, Albert Majura, Naike Nathaniel, Julietha Tibyesiga, Loveness Kimaro, Judith Kokuleba, Ngusa Kalambo, Constancia F Luyenga, Monica Mtei, Jackline Ngowi, Edson B Jeremiah, Witness Simon, Erick Kazoka, Chrispin Mgute.

## Notes

### Competing Interest Statement

The authors have declared no competing interest.

### Funding Statement

This research work was supported by grant from UNICEF (MA). The funders had no role in study design, data collection and analysis, decision to publish, or preparation of the manuscript.

### Author Declarations

Ethical approval was obtained from the Research and Publication Committee of the Muhimbili University of Health and Allied Sciences (MUHAS-REC-08-2021-839). Permission to collect data in Regions and Councils was sought from the President’s Office Regional Administration and Local Government, Ministry of Health Community, Development, Gender, Elderly and Children (MoHCDGEC), Regional Secretariat (RS) and Local Government Authorities (LGAs).

## References

1. Dereje N, Tesfaye A, Tamene B, Alemeshet D, Abe H, Tesfa N, et al. COVID-19 vaccine hesitancy in Addis Ababa, Ethiopia: a mixed-method study. BMJ Open. 2022 May 30;12(5):e052432. doi: 10.1136/bmjopen-2021-052432. PMID: 35636790; PMCID: PMC9152622.

2. COVID-19 dashboard [Internet]. [cited 2022 Jan 19]. Available from: https://www.gavi.org/covid19/dashboard

3. United Republic of Tanzania: WHO Coronavirus Disease (COVID-19) Dashboard With Vaccination Data | WHO Coronavirus (COVID-19) Dashboard With Vaccination Data [Internet]. [cited 2022 Jan 19]. Available from: https://covid19.who.int/region/afro/country/tz

4. Angelo AT, Alemayehu DS, Dachew AM. Health care workers intention to accept COVID-19 vaccine and associated factors in southwestern Ethiopia, 2021. PLoS One. 2021 Sep 3;16(9):e0257109. doi: 10.1371/journal.pone.0257109. PMID: 34478470; PMCID: PMC8415602.

5. WHO SAGE Roadmap For Prioritizing Uses Of COVID-19 Vaccines In The Context Of Limited Supply [Internet]. [cited 2022 Jan 19]. Available from: https://www.who.int/publications/i/item/who-sage-roadmap-for-prioritizing-uses-of-covid-19-vaccines-in-the-context-of-limited-supply

6. Tanzania Publishes First COVID-19 Data in Over a Year. [cited 2022 Jan 20]. Available from: https://www.voanews.com/a/africa_tanzania-publishes-first-covid-19-data-over-year/6207608.html

7. WHO. Ten threats to global health in Air pollution and climate change Noncommunicable diseases. 2019;1–9. Available from: https://www.who.int/news-room/spotlight/ten-threats-to-global-health-in-2019

8. Liu Q, Luo D, Haase JE, Guo Q, Wang XQ, Liu S, et al. The experiences of health-care providers during the COVID-19 crisis in China: a qualitative study. Lancet Glob Health. 2020 Jun;8(6):e790–e798. doi: 10.1016/S2214-109X(20)30204-7. Epub 2020 Apr 29. PMID: 32573443; PMCID PMC7190296.

9. Ferrer R, Klein WM. Risk perceptions and health behavior. Curr Opin Psychol. 2015 Oct 1;5:85–89. doi: 10.1016/j.copsyc.2015.03.012. PMID: 26258160; PMCID PMC4525709.

10. Barrett ES, Horton DB, Roy J, Gennaro ML, Brooks A, Tischfield J, et al. Prevalence of SARS-CoV-2 infection in previously undiagnosed health care workers at the onset of the U.S. COVID-19 epidemic. medRxiv [Preprint]. 2020 Apr 24:2020.04.20.20072470. doi: 10.1101/2020.04.20.20072470. Update in: BMC Infect Dis. 2020 Nov 16;20(1):853. PMID: 32511600; PMCID PMC7276027.

11. Sikkema RS, Pas SD, Nieuwenhuijse DF, O’Toole Á, Verweij J, van der Linden A, et al. COVID-19 in health-care workers in three hospitals in the south of the Netherlands: a cross-sectional study. Lancet Infect Dis. 2020 Nov;20(11):1273-1280. doi: 10.1016/S1473-3099(20)30527-2. Epub 2020 Jul 2. Erratum in: Lancet Infect Dis. 2020 Sep;20(9):e215. PMID: 32622380; PMCID PMC7332281.

12. Liao Q, Wu P, Wing Tak Lam W, Cowling BJ, Fielding R. Trajectories of public psycho-behavioural responses relating to influenza A(H7N9) over the winter of 2014-15 in Hong Kong. Psychol Health. 2019 Feb;34(2):162–180. doi: 10.1080/08870446.2018.1515436. Epub 2018 Nov 15. PMID: 30430862.

13. Deressa W, Worku A, Abebe W, Gizaw M, Amogne W. Risk perceptions and preventive practices of COVID-19 among healthcare professionals in public hospitals in Addis Ababa, Ethiopia. PLoS One. 2021 Jun 25;16(6):e0242471. doi: 10.1371/journal.pone.0242471. PMID: 34170910; PMCID PMC8232450.

14. Graffigna G, Palamenghi L, Boccia S, Barello S. Relationship between Citizens’ Health Engagement and Intention to Take the COVID-19 Vaccine in Italy: A Mediation Analysis. Vaccines (Basel). 2020 Oct 1;8(4):576. doi: 10.3390/vaccines8040576. PMID: 33019663; PMCID PMC7711984.

15. Ba MF, Faye A, Kane B, Diallo AI, Junot A, Gaye I, et al. Factors associated with COVID-19 vaccine hesitancy in Senegal: A mixed study. Hum Vaccin Immunother. 2022 Nov 30;18(5):2060020. doi: 10.1080/21645515.2022.2060020. Epub 2022 May 11. PMID: 35543616.

16. Kanu S, James PB, Bah AJ, Kabba JA, Kamara MS, Williams CEE, et al. Healthcare Workers’ Knowledge, Attitude, Practice and Perceived Health Facility Preparedness Regarding COVID-19 in Sierra Leone. J Multidiscip Healthc. 2021 Jan 11;14:67–80. doi: 10.2147/JMDH.S287156. PMID: 33469299; PMCID PMC7810694.

17. Fares S, Elmnyer MM, Mohamed SS, Elsayed R. COVID-19 Vaccination Perception and Attitude among Healthcare Workers in Egypt. J Prim Care Community Health. 2021 Jan-Dec;12:21501327211013303. doi: 10.1177/21501327211013303. PMID: 33913365; PMCID PMC8111272.

18. Detoc M, Bruel S, Frappe P, Tardy B, Botelho-Nevers E, Gagneux-Brunon A. Intention to participate in a COVID-19 vaccine clinical trial and to get vaccinated against COVID-19 in France during the pandemic. Vaccine. 2020 Oct 21;38(45):7002–7006. doi: 10.1016/j.vaccine.2020.09.041. Epub 2020 Sep 17. PMID: 32988688; PMCID PMC7498238.

19. Seale H, Heywood AE, Leask J, Sheel M, Durrheim DN, Bolsewicz K, et al. Examining Australian public perceptions and behaviors towards a future COVID-19 vaccine. BMC Infect Dis. 2021 Jan 28;21(1):120. doi: 10.1186/s12879-021-05833-1. PMID: 33509104; PMCID PMC7840792.

20. Sallam M, Dababseh D, Eid H, Al-Mahzoum K, Al-Haidar A, Taim D, et al. High Rates of COVID-19 Vaccine Hesitancy and Its Association with Conspiracy Beliefs: A Study in Jordan and Kuwait among Other Arab Countries. Vaccines (Basel). 2021 Jan 12;9(1):42. doi: 10.3390/vaccines9010042. PMID: 33445581; PMCID PMC7826844.

21. Ciardi F, Menon V, Jensen JL, Shariff MA, Pillai A, Venugopal U, et al. Knowledge, Attitudes and Perceptions of COVID-19 Vaccination among Healthcare Workers of an Inner-City Hospital in New York. Vaccines (Basel). 2021 May 17;9(5):516. doi: 10.3390/vaccines9050516. PMID: 34067743; PMCID PMC8156250.

22. Creswell JW, Poth CN. Qualitative Inquiry and Research Design: Choosing among five approaches. SAGE Publications; 2016.

23. Braun V, Clarke V. What can “thematic analysis” offer health and wellbeing researchers? Int J Qual Stud Health Well-being. 2014 Oct 16;9:26152. doi: 10.3402/qhw.v9.26152. PMID: 25326092; PMCID PMC4201665.

24. Sarah Dryhurst, Claudia R. Schneider, John Kerr, Alexandra L. J. Freeman, Gabriel Recchia, Anne Marthe van der Bles, David Spiegelhalter & Sander van der Linden. Risk perceptions of COVID-19 around the world, Journal of Risk Research. 2020; 23:7-8, 994-1006, DOI: 10.1080/13669877.2020.1758193

25. Abdel Wahed WY, Hefzy EM, Ahmed MI, Hamed NS. Assessment of Knowledge, Attitudes, and Perception of Health Care Workers Regarding COVID-19, A Cross-Sectional Study from Egypt. J Community Health. 2020 Dec;45(6):1242–1251. doi: 10.1007/s10900-020-00882-0. PMID: 32638199; PMCID PMC7340762.

26. Solís Arce JS, Warren SS, Meriggi NF, Scacco A, McMurry N, Voors M, et al. COVID-19 vaccine acceptance and hesitancy in low-and middle-income countries. Nat Med. 2021 Aug;27(8):1385–1394. doi: 10.1038/s41591-021-01454-y. Epub 2021 Jul 16. PMID: 34272499; PMCID PMC8363502.

27. Anjorin AA, Odetokun IA, Abioye AI, Elnadi H, Umoren MV, Damaris BF, et al. Will Africans take COVID-19 vaccination? PLoS One. 2021 Dec 1;16(12):e0260575. doi: 10.1371/journal.pone.0260575. PMID: 34851998; PMCID PMC8635331.

28. Karlsson LC, Soveri A, Lewandowsky S, Karlsson L, Karlsson H, Nolvi S, et al. Fearing the disease or the vaccine: The case of COVID-19. Pers Individ Dif. 2021 Apr;172:110590. doi: 10.1016/j.paid.2020.110590. Epub 2020 Dec 14. PMID: 33518869; PMCID PMC7832025.

29. Anjorin AA, Abioye AI, Asowata OE, Soipe A, Kazeem MI, Adesanya IO, et al. Comorbidities and the COVID-19 pandemic dynamics in Africa. Trop Med Int Health. 2021 Jan;26(1):2–13. doi: 10.1111/tmi.13504. Epub 2020 Oct 23. PMID: 33012053; PMCID PMC7675305.

30. Abayomi A, Osibogun A, Kanma-Okafor O, Idris J, Bowale A, Wright O, Adebayo B, et al. Morbidity and mortality outcomes of COVID-19 patients with and without hypertension in Lagos, Nigeria: a retrospective cohort study. Glob Health Res Policy. 2021 Jul 29;6(1):26. doi: 10.1186/s41256-021-00210-6. Erratum in: Glob Health Res Policy. 2021 Aug 13;6(1):28. PMID: 34325747; PMCID PMC8319704.

31. Diop BZ, Ngom M, Pougué Biyong C, Pougué Biyong JN. The relatively young and rural population may limit the spread and severity of COVID-19 in Africa: a modelling study. BMJ Glob Health. 2020 May;5(5):e002699. doi: 10.1136/bmjgh-2020-002699. Erratum in: BMJ Glob Health. 2020 Jul;5(7): PMID: 32451367; PMCID PMC7252974.

32. Finucane, M.L., Alhakami, A., Slovic, P. and Johnson, S.M. The affect heuristic in judgments of risks and benefits. J. Behav. Decis. Making. 2000; 13: 1–17. https://doi.org/10.1002/(SICI)1099-0771(200001/03)13:1<1::AID-BDM333>3.0.CO;2-S

